# Avoided and avoidable deaths with the use of COVID-19 convalescent plasma in Italy during the first two years of pandemic

**DOI:** 10.1101/2024.08.12.24311864

**Authors:** Massimo Franchini, Arturo Casadevall, Quigly Dragotakes, Daniele Focosi

## Abstract

Italy was the first western country to be hit by the COVID-19 pandemic and suffered nearly 200,000 deaths so far during the four years of the pandemic. In March 2020, Italy first deployed COVID-19 convalescent plasma (CCP) to treat hospitalized patients. Despite this initial effort, the proportion of COVID-19 patients treated with CCP during the first two years of the pandemic (2020-2021) was very low (approximately 2% of individuals hospitalized for COVID-19). In this study, we estimated the number of actual inpatient lives saved by CCP treatment in Italy using national mortality data, and CCP mortality reduction data from meta-analyses of randomized controlled trials and real-world data. We also estimated the potential number of lives saved if CCP had been deployed to 100% of hospitalized patients or used in 15% to 75% of outpatients. According to these models, CCP usage in 2020-2021 saved between 385-1304 lives, but this number would have increased to 17,751-60,079 if 100% of inpatients had been transfused with CCP. Similarly, broader (15-75%) usage in outpatients could have prevented 21,187-190,689 hospitalizations (desaturating hospitals) and 6,144-81,926 deaths. These data have important implications for convalescent plasma use in future infectious disease emergencies.

## Introduction

Since 2019, the SARS-CoV-2 pandemic has generated an unprecedented health and social emergency, with the world largely unprepared for a such new viral disease [1]. The first therapeutic weapon used to fight the pathogen was plasma from recovered people (COVID-19 convalescent plasma, CCP), based on historical efficacy and the *in vitro* demonstration that it contained antibodies able to neutralize viral infection [2-4]. After more than 4 years of the pandemic and nearly 50 randomized controlled trials (RCTs), the role of CCP has been finally clarified: when transfused at high titer (> 180) and early (within 7 days from symptom onset), CCP is safe and effective in both ambulatory and hospitalized patients [5-8]. Today, with most of the population having endogenous antibody from vaccination and/or SARS-CoV-2 infection the primary use of CCP is in immunocompromised patients, who are not able to produce and adequate antibody response against the virus [9,10]. In addition, due to the frequent mutations that render SARS-CoV-2 resistant to the authorized anti-Spike monoclonal antibodies, CCP is currently the only effective antibody-based antiviral therapy, being able to follow closely the natural evolution of the virus.

In a recently pre-published study from the US, where over 500,000 hospitalized COVID-19 patients were treated with CCP during the first year of the pandemic, the investigators estimated that the extensive use of CCP in hospitalized COVID-19 patients would have saved between 16,476 and 66,296 lives, while its extensive outpatient use would have prevented 1,136,880 hospitalizations and saved 215,195 lives [11].

The aim of this study was to replicate such analysis using Italian statistics and efficacy numbers, to estimate the potential impact of CCP use at reducing mortality from SARS-COV-2 in Italy during the first two years (2020-2021) of the pandemic.

## Methods

To estimate the potential number of avoided and avoidable deaths with CCP use during the years 2020 and 2021 of the pandemic, the following mathematical model was developed.

### CCP units transfused and patients treated

According to the official data from the Italian Ministry of Health [12], in the year 2020 6,952 plasmapheresis procedures were performed, followed by 9,301 in the year 20219. Each CCP plasmapheresis had a 600 ml volume. The number of CCP units (each ranging from 200 to 300 ml) derived from these donations were 13,731 for 2020 and 14,558 for 2021. Overall, 28,289 CCP units were collected. Of these, 1773 were discarded for reasons linked to health, technical issues and quality control. Overall, 20,438 CCP units were transfused (6,912 in 2020 and 13,526 in 2021). According to the data from Italian RCTs [13,14] and real-life studies [15,16], each patient received a median of 2 CCP units. Then a total of 10,219 COVID-19 patients (3456 in the year 2020 and 6763 in the year 2021) were transfused each with 2 CCP units during the years 2020 and 2021.

### Number of COVID-19-related hospitalizations and deaths

We calculated the number of hospitalizations for COVID-19 and in-hospital COVID-19 related deaths in Italy during the years 2020 and 2021. The data were extracted from the daily report of the Italian Ministry of Health [17,18]. According to these data, during the year 2020 the number of deaths/hospitalizations was 77,871/239,963 (32.4% mortality) while in the year 2021 the number of deaths/hospitalizations was 58,627/230,874 (25.4% mortality). Overall, during the two years period there were 136,498 deaths among 470,837 hospitalized patients (29.0% mortality).

### CCP mortality reduction percentages

We made 3 different estimates of the CCP mortality relative risk reduction (RRR) The first estimate was based on the Italian RCT TSUNAMI [13], who found a 23% RRR in the 30-day mortality (6.1% in the CCP group versus 7.9% in controls). The second estimate used the real-world data from the largest Italian registry (Veneto region), reporting 30-day mortality RRR of 44% during the period February 2020 and June 2021 (14% in the CCP group versus 25% in controls) [15]. We note that this 44% mortality RRR for CCP treated individuals is in the range of the 47% effect reported in a large propensity-score matched real-world study from the USA [19]. The third estimate was based on a systematic review and meta-analysis of all CCP controlled studies performed in 2022 (including 39 RCTs with 21,529 participants and 70 controlled cohort studies with 50,160 participants), which estimated that CCP reduced mortality by 13% among hospitalized patients [6].

### Estimating actual and potential lives saved by CCP use in hospitalized patients

Actual lives saved by CCP were calculated using the three separate estimates of mortality benefit conferred by CCP in hospitalized patients detailed above (i.e., 23%, 44% and 13%) [6,13,15]. The formula used to calculate the number of lives saved by CCP in comparison to a situation where CCP was never used was:

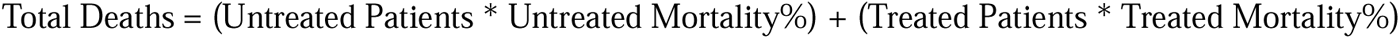

We then calculated the lives saved as the difference between the above and (Admissions * Untreated Mortality) where the comparator was the absence of CCP treatment, using the three-mortality reduction estimated described above from trials and real-world data (*i.e.* 23%, 44%, and 13%).

In addition to the real-life numbers, we estimated the number of lives that would have been saved if CCP had been administered to 100% of hospitalized patients (potential lives saved), again using the three measures of efficacy in reducing mortality described above. The formula used was similar to that previously reported, except for the assumption that all hospitalized patients would have received CCP.

### Estimating potential lives saved if CCP had been deployed for outpatient use

Given the greater efficacy of CCP when used early in the course of disease [7], it is reasonable that outpatient use could have saved even more lives than inpatient use. A RCT of CCP outpatient efficacy early in the pandemic reported a 48% RRR in progression to severe illness likely to lead to hospitalization among elderly patients [20]. Subsequently, a large RCT of CCP outpatient use reported a 54.3% RRR in progression to hospitalization [21]. Consequently, we estimated the potential lives saved by outpatient use based on outpatient CCP efficacy data obtained during the pandemic. When CCP was given in the first 5 days of symptoms, RRR in progression to hospitalization rose to 79.9%, which is similar to that of anti-Spike monoclonal antibodies [7]. A more conservative figure of 30% for outpatient CCP emerges from a meta-analysis of 5 RCTs including international trials [22]. We used all these 3 estimates (i.e., 30%, 54% and 80%) in our analysis. In addition, since the great majority of COVID-19 patients died in the hospitals, it was possible to estimate the potential number of lives saved by deployment of outpatient CCP according to the following formula:

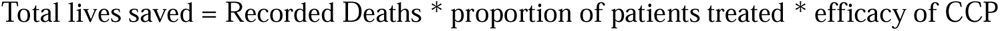

The prevalence of use of CCP in COVID-19 outpatients at high risk for disease progression and hospitalization was estimated from the actual use of anti-Spike monoclonal antibodies in the US (15%; Italian data are not available) [23] or from a hypothetical major national effort to deploy outpatient CCP use (75%).

## Results

Table 1 reports the data used to calculate the real and potential number of lives saved by in-hospital CCP use (data retrieved from the Italian Ministry of Health) [18]. Overall, 10,219 hospitalized COVID-19 patients received 2 units of CCP during the years 2020 and 2021 (3,456 in the year 2020 and 6,763 in the year 2021), representing the 2.2% (10,219/470,837) of the total number of patients admitted for COVID-19 during the two-year period. Overall, 29.0% of COVID-19 hospitalized patients died for COVID-19 (32.4% [77,871/239,963] in the year 2020 and 25.4 [58,627/230,874] in the year 2021).

**Table 1.** Input data used for the estimation of live saved from CCP in Italy.

| **Year** | **2020** | **2021** | **Total (2020-2021)** |
| --- | --- | --- | --- |
| **COVID-19 patients treated (n)<sup>1</sup>** | 3,456 | 6,763 | 10,219 |
| **Hospitalizations for COVID-19** | 239,963 | 230,874 | 470,837 |
| **COVID-related deaths** | 77,871 | 58,627 | 136,498 |
| **COVID-related deaths /**<br>**hospitalizations (%)** | 32.4 | 25.4 | 29.0 |

<sup>1</sup>Each patient received two CCP units.

Table 2 and 3 report the estimates of the actual number of lives saved by in-hospital CCP use during the years 2020 and 2021, respectively. The number of lives saved by CCP use ranged from 62 in 2020 (using the 1.8% mortality RRR provided by reference 13) to 879 in 2021 (using the 13% mortality RRR provided by reference 6). If CCP would have been transfused to the whole population of hospitalized patients, the lives saved would have increased to a range from a minimum of 7,623 (year 2021, 13% mortality reduction model) up to a maximum of34,209 (year 2020, 44% mortality reduction model).

**Table 2.**
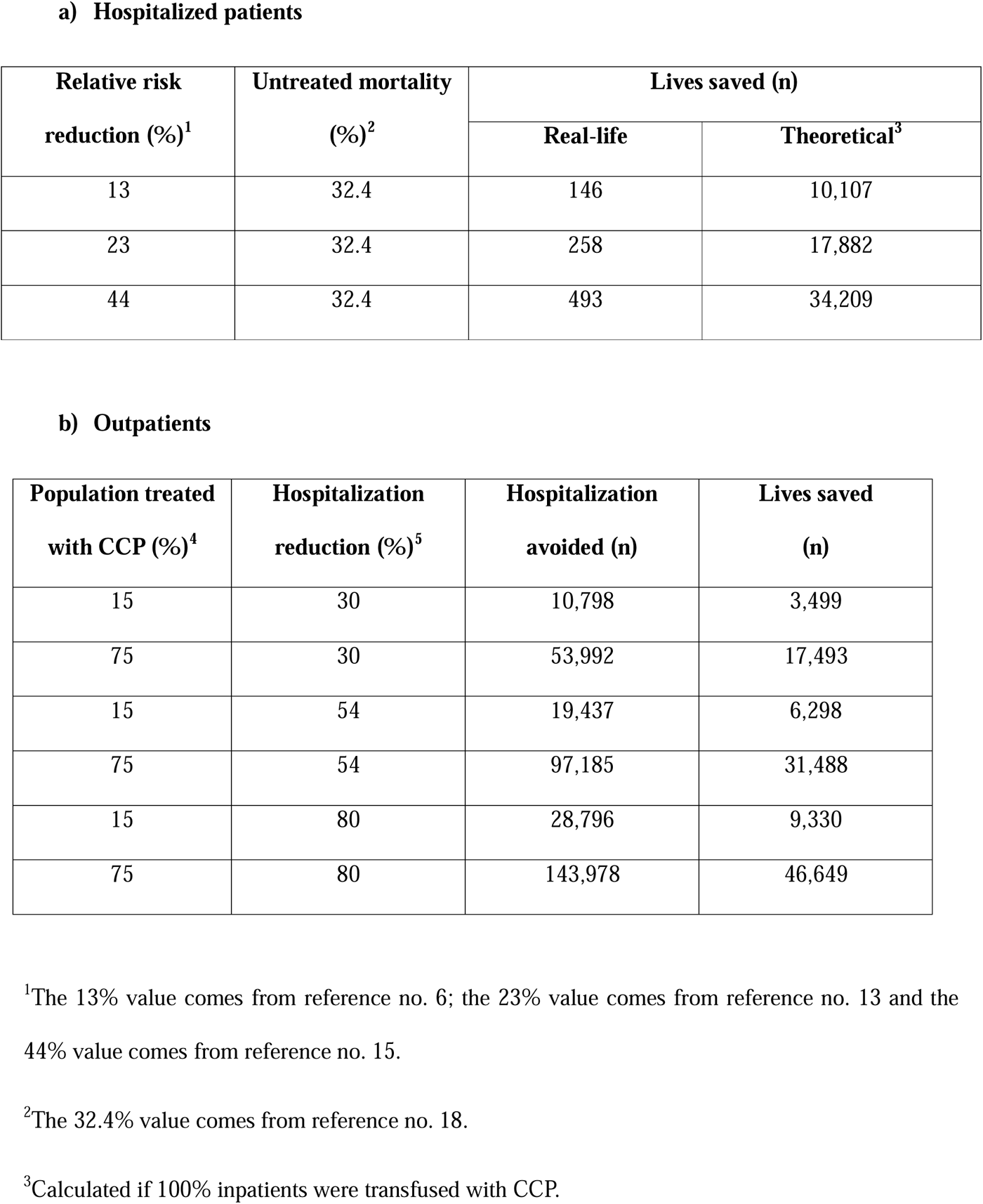
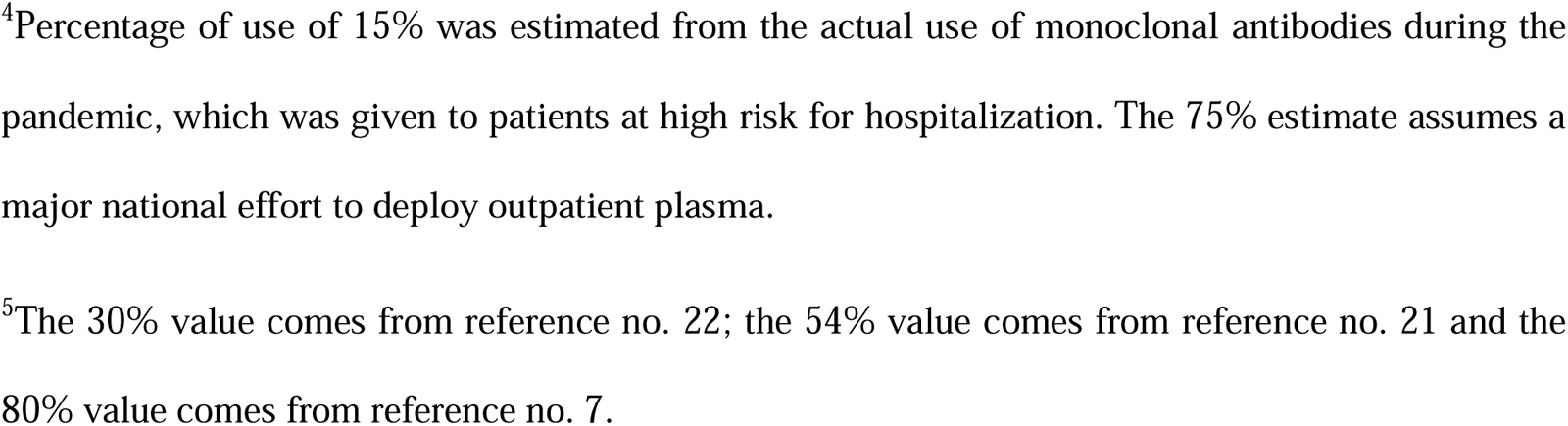
Estimates of actual and potential lives saved from the deployment of CCP in Italy, year 2020.

**a) Hospitalized patients**
| **Relative risk reduction (%)<sup>1</sup>** | **Untreated mortality (%)<sup>2</sup>** | **Lives saved (n)** |  |
| --- | --- | --- | --- |
|  |  | **Real-life** | **Theoretical<sup>3</sup>** |
| 13 | 32.4 | 146 | 10,107 |
| 23 | 32.4 | 258 | 17,882 |
| 44 | 32.4 | 493 | 34,209 |

**b) Outpatients**
| **Population treated with CCP (%)<sup>4</sup>** | **Hospitalization reduction (%)<sup>5</sup>** | **Hospitalization avoided (n)** | **Lives saved (n)** |
| --- | --- | --- | --- |
| 15 | 30 | 10,798 | 3,499 |
| 75 | 30 | 53,992 | 17,493 |
| 15 | 54 | 19,437 | 6,298 |
| 75 | 54 | 97,185 | 31,488 |
| 15 | 80 | 28,796 | 9,330 |
| 75 | 80 | 143,978 | 46,649 |

<sup>1</sup>The 13% value comes from reference no. 6; the 23% value comes from reference no. 13 and the 44% value comes from reference no. 15.
<sup>2</sup>The 32.4% value comes from reference no. 18.
<sup>3</sup>Calculated if 100% inpatients were transfused with CCP.
<sup>4</sup>Percentage of use of 15% was estimated from the actual use of monoclonal antibodies during the pandemic, which was given to patients at high risk for hospitalization. The 75% estimate assumes a major national effort to deploy outpatient plasma.
<sup>5</sup>The 30% value comes from reference no. 22; the 54% value comes from reference no. 21 and the 80% value comes from reference no. 7.

Tables 2 and 3 also list the estimate of hospitalizations prevented and lives saved in the outpatient setting. According to this model, the number of hospitalizations avoided ranged from a conservative minimum of 10,798 (15% of high-risk COVID-19 outpatients transfused, and 30% hospitalization RRR) up to a maximum 143,978 (75% of high-risk COVID-19 outpatients treated, and 80% hospitalization RRR) in the year 2020, and from 10,389 (15% of high-risk COVID-19 outpatients treated and 30% hospitalization RRR) to 138,524 (75% of high-risk COVID-19 outpatients treated and 80% hospitalization RRR) in the year 2021. Accordingly, the number of lives saved ranged from 2,700 (15% of high-risk COVID-19 outpatients treated and 30% RRR in hospitalization) to 35,994 (75% of high-risk COVID-19 outpatients treated and 80% RRR in hospitalization) in the year 2020 and from 2,597 (15% of high-risk COVID-19 outpatients treated and 30% hospitalization RRR) to 34,631 (75% of high-risk COVID-19 outpatients treated and 80% hospitalization RRR) in the year 2021.

**Table 3.** Estimates of actual and potential lives saved from the deployment of CCP in Italy, year 2021.

**a) Hospitalized patients**
| **Relative risk reduction (%)<sup>1</sup>** | **Untreated mortality (%)<sup>2</sup>** | **Lives saved (n)** |  |
| --- | --- | --- | --- |
|  |  | **Real-life** | **Theoretical<sup>3</sup>** |
| 13 | 25.4 | 223 | 7,623 |
| 23 | 25.4 | 395 | 13,488 |
| 44 | 25.4 | 756 | 25,802 |

**b) Outpatients**
| **Population treated with CCP (%)<sup>4</sup>** | **Hospitalization reduction (%)<sup>5</sup>** | **Hospitalization avoided (n)** | **Lives saved (n)** |
| --- | --- | --- | --- |
| 15 | 30 | 10,389 | 2,639 |
| 75 | 30 | 51,947 | 13,194 |
| 15 | 54 | 18,701 | 4,750 |
| 75 | 54 | 93,504 | 23,750 |
| 15 | 80 | 27,705 | 7,037 |
| 75 | 80 | 138,524 | 35,185 |

<sup>1</sup>The 13% value comes from reference no. 6; the 23% value comes from reference no. 13 and the 44% value comes from reference no. 15.
<sup>2</sup>The 25.4% value comes from reference no. 18.
<sup>3</sup>Calculated if 100% inpatients were transfused with CCP.
<sup>4</sup>Percentage of use of 15% was estimated from the actual use of monoclonal antibodies during the pandemic, which was given to patients at high risk for hospitalization. The 75% estimate assumes a major national effort to deploy outpatient plasma.
<sup>5</sup>The 30% value comes from reference no. 21; the 54% value comes from reference no. 21 and the 80% value comes from reference no. 7.

Considering the two-year period (Table 4), the actual real-life and 100% treated theoretically saved lives by CCP in-hospital use ranged from 385 to 1,304 (13% to 44% mortality RRR) and from 17,751 to 60,079 (13% to 44% mortality RRR), respectively. During the same period, the number of hospitalizations avoided, and lives saved by CCP outpatient use ranged from 21,188 to 282,502 (15% to 75% of outpatients treated and 30% to 80% hospitalization RRR) and from 6,144 to 81,926 (15% to 75% of outpatients treated and 30% to 80% hospitalization RRR), respectively.

**Table 4.** Estimates of actual and potential lives saved from the deployment of CCP in Italy, years 2020-2021.

**a) Hospitalized patients**
| **Relative risk reduction (%)<sup>1</sup>** | **Untreated mortality (%)<sup>2</sup>** | **Lives saved (n)** |  |
| --- | --- | --- | --- |
|  |  | **Real-life** | **Theoretical<sup>3</sup>** |
| 13 | 29 | 385 | 17,751 |
| 23 | 29 | 682 | 31,405 |
| 44 | 29 | 1,304 | 60,079 |

**b) Outpatients**
| **Population treated with CCP (%)<sup>4</sup>** | **Hospitalization reduction (%)<sup>5</sup>** | **Hospitalization avoided (n)** | **Lives saved (n)** |
| --- | --- | --- | --- |
| 15 | 30 | 21,187 | 6,144 |
| 75 | 30 | 105,939 | 30,722 |
| 15 | 54 | 38,138 | 11,060 |
| 75 | 54 | 190,689 | 55,300 |
| 15 | 80 | 56,501 | 16,385 |
| 75 | 80 | 282,502 | 81,926 |

<sup>1</sup>The 13% value comes from reference no. 6; the 23% value comes from reference no. 13 and the 44% value comes from reference no. 15.
<sup>2</sup>The 29% value comes from reference no. 18.
<sup>3</sup>Calculated if 100% inpatients were transfused with CCP.
<sup>4</sup>Percentage of use of 15% was estimated from the actual use of monoclonal antibodies during the pandemic, which was given to patients at high risk for hospitalization. The 75% estimate assumes a major national effort to deploy outpatient plasma.
<sup>5</sup>The 30% value comes from reference no. 21; the 54% value comes from reference no. 21 and the 80% value comes from reference no. 7.

## Discussion

Although Italy was the first western country to be hit by COVID-19 pandemic and the first to use CCP in inpatients, only a negligible proportion (approximately 2%) of patients hospitalized for COVID-19 received this potentially life-saving biological antiviral therapy during the first two years (2020 and 2021) of the pandemic. This disappointing finding contrasts profoundly with data from the US, which by the Fall of 2020 as many as 40% of hospitalized COVID-19 patients were being treated with CCP [24]. The reasons for such a difference were multiple. First, the publication of the early Italian positive experience on CCP use in hospitalized COVID-19 patients [3] poorly stimulated further CCP collection and transfusion in Italy, and was received with caution by the Italian scientific community, in particular by the experts within the technical-scientific committee of the Italian Ministry of Health, who recommended caution with in-hospital CCP use until the Italian RCT, named TSUNAMI, had been published. The RCT was published in late 2021 [13], but its negative results were anticipated in the form of a succinct press release in early 2021 [25], which had the consequence of stopping the already low use of CCP in Italian hospitals. In addition, the Italian Society of Transfusion Medicine and Immunohematology (SIMTI) itself recommended against the administration of CCP in hospitalized COVID-19 patients [26]. However, upon careful reading, the results of the Italian TSUNAMI RCT were not completely negative, but showed a clear trend towards efficacy (P = 0.06) of CCP versus standard therapy in the subgroup of patients with milder (and thus earlier) disease. Today we know from a compilation of RCT that this population can benefit from CCP [8], and that the TSUNAMI trend was almost certainly a real effect. In retrospect, this finding is not surprising for as it matched exactly what happened with other RCTs of anti-Spike monoclonal antibodies and CCP in outpatient setting [7]. Ideally, the TSUNAMI observation should have provided the basis for the design of a more modern RCT focusing on treatment of earlier phases (i.e., outpatients) of COVID-19 with CCP. Unfortunately, this did not happen, and national authorities did not support any additional investigation on CCP. Consequently, CCP has remained in Italy an experimental biological product requiring regulatory authorizations (which delayed its transfusion) before its restricted in-hospital use. The Italian experience was very different from that in the USA, where CCP was first authorized for in-hospital use (under an Expanded Access Program [EAP] in April 2020 and under Emergency Use Authorization [EUA] in August 2020] and then for outpatient use in immunosuppressed patients (December 2021). The deployment of CCP in the USA saved a significant number of lives during the pandemic, positively impacting the overall COVID-related mortality rates, which were lower in those geographical areas where CCP was used more extensively [24]. The fact that CCP has remained an experimental biological product in Italy has also prevented its outpatient use, which would have reduced hospitalizations saving further lives and desaturating the heavily crowded hospitals. It is noteworthy that in the USA the FDA has issued guidance to industry for the licensing of CCP in July 2024 [27], which attests to regulatory acceptance of its safety and efficacy and the need for continued supply of this product given its usefulness in immunocompromised individuals. As a consequence, the number of COVID-19 patients treated with CCP and the number of lives saved in Italy during the first two years of pandemic were much smaller than it could have been, while a widespread early use of CCP would certainly have contributed to reduce the nearly 200,000 deaths recorded during the four years of the pandemic [28].

But what is the goal of this study? It is not so much to criticize the efforts during the pandemic but to highlight the real and potential effectiveness that CCP therapy has had in the fight against COVID-19 and to take this information into consideration in the event of future infectious emergencies. Under such scenarios, we recommend that convalescent plasma be deployed on the basis of its historical efficacy (including COVID-19), using registries that allow the collection of efficacy data that can be exploited to inform the design of RCTs. Registries allow the therapy to be immediately available to the citizens and can provide critical information on safety, dose and timing to inform RCT design [29,30]. The argument that registries preclude the completion of RCTs is not viable given that the USA completed at least 5 RCTs while CCP was available under FDA EUA. We also encourage researchers from other nations to run similar nation-wide analyses, to obtain information that can encourage governments to include convalescent plasma therapy in their future pandemic preparedness plans.

## Data Availability

This manuscript generated no new dataset.

## Abbreviations

ARR: absolute risk reduction
CCP: COVID-19 convalescent plasma
RRR: relative risk reduction.

## Notes

### Competing Interest Statement

The authors have declared no competing interest.

### Funding Statement

This study did not receive any funding

